# Spatial frequency discrimination in patients with schizophrenia spectrum and bipolar disorders: Evidence of early visual processing deficits and associations with intellectual abilities

**DOI:** 10.1101/2021.06.28.21259309

**Authors:** Aili R. Løchen, Knut K. Kolskår, Ann-Marie G. de Lange, Markus H. Sneve, Beathe Haatveit, Trine V. Lagerberg, Torill Ueland, Ingrid Melle, Ole A. Andreassen, Lars T. Westlye, Dag Alnæs

**Author notes:** Corresponding authors: Aili R. Løchen & Dag Alnæs, *Oslo University Hospital, PO Box 4956 Nydalen, 0424 Oslo, Norway,.

## Abstract

**Objective:** Low-level sensory disruption is hypothesized as a precursor to clinical and cognitive symptoms in severe mental disorders. We compared visual discrimination performance in patients with schizophrenia spectrum disorder or bipolar disorder with healthy controls, and investigated associations with clinical symptoms and IQ.

**Methods:** Patients with schizophrenia spectrum disorder (n=32), bipolar disorder (n=55) and healthy controls (n=152) completed a computerized visual discrimination task. Participants responded whether the latter of two consecutive grids had higher or lower spatial frequency, and discrimination thresholds were estimated using an adaptive maximum likelihood procedure. Case-control differences in threshold were assessed using linear regression, F-test and post-hoc pair-wise comparisons. Linear models were used to test for associations between visual discrimination threshold and psychotic symptoms derived from the PANSS and IQ assessed using the Matrix Reasoning and Vocabulary subtests from the Wechsler Abbreviated Scale of Intelligence (WASI).

**Results:** Robust regression revealed a significant main effect of diagnosis on discrimination threshold (robust F=6.76, p=.001). Post-hoc comparisons revealed that patients with a schizophrenia spectrum disorder (mean=14%, SD=0.08) had higher thresholds compared to healthy controls (mean=10.8%, SD = 0.07, β = 0.35, t=3.4, p=0.002), as did patients with bipolar disorder (12.23%, SD=0.07, β= 0.21, t=2.42, p=0.04). There was no significant difference between bipolar disorder and schizophrenia (β=-0.14, t=-1.2, p=0.45). Linear models revealed negative associations between IQ and threshold across all participants when controlling for diagnostic group (β = -0.3, t=-3.43, p=0.0007). This association was found within healthy controls (t=-3.72, p=.0003) and patients with bipolar disorder (t=-2.53, p=.015), and no significant group by IQ interaction on threshold (F=0.044, p=.97). There were no significant associations between PANSS domain scores and discrimination threshold.

**Conclusion:** Patients with schizophrenia spectrum or bipolar disorders exhibited higher visual discrimination thresholds than healthy controls, supporting early visual deficits among patients with severe mental illness. Discrimination threshold was negatively associated with IQ among healthy controls and bipolar disorder patients. These findings elucidate perception-related disease mechanisms in severe mental illness, which warrants replication in independent samples.

## Introduction

Schizophrenia and bipolar disorder are severe mental disorders characterized by perceptual, cognitive and emotional symptoms (Rowland & Marwaha, 2018). Perceptual disruptions across several domains are defining features of schizophrenia, but are also frequently reported in bipolar disorder (O’Bryan, Brenner, Hetrick, & O’Donnell, 2014), both in manic and depressive phases (Parker, Paterson, Romano, & Graham, 2017). Previous studies of patients with schizophrenia have reported disruptions in speech (Kuperberg, 2010), object (Gabrovska, Laws, Sinclair, & McKenna, 2003) and face (Megreya, 2016) perception, as well as in basic sensory and perceptual processes (Javitt, 2009a), including tone-matching, contrast sensitivity, and spatial frequency discrimination.

Low-level sensory and perceptual mechanisms enable the organism to detect, discriminate and categorize basic sensory features, such as pitch and tone discrimination in the auditory modality, and orientation, spatial frequency and colors in the visual domain (Eggermont & Ponton, 2002; Kravitz, Saleem, Baker, Ungerleider, & Mishkin, 2013; Riesenhuber & Poggio, 1999). These functions reflect partly automated “bottom-up” cortical processing, and constitute the early stages of the sensory and perceptual streams in the visual processing hierarchy (Mechelli, Price, Friston, & Ishai, 2004). These building blocks give rise to higher-level perceptual abstractions such as objects, faces, location and movement (Peissig & Tarr, 2007; Perry & Fallah, 2014). Importantly, this complex integrative process has been shown to also involve higher-level cognition or “top-down” executive functions such as decision making, memory, planning, (Gilbert & Li, 2013; Vetter & Newen, 2014). Disruptions in these early sensory and perceptual pathways are hypothesized to constitute precursors to the abnormal higher-level abstract perceptual experiences (e.g. hallucinations) and beliefs (e.g. delusions) often reported by patients with severe mental illness (Conde, Gonçalves, & Pinheiro, 2016; Phillipson & Harris, 1985; Pinheiro et al., 2017).

The relevance of visual discrimination performance and sensory impairments for mental illness is supported by several lines of evidence. A prospective study of children of mothers with schizophrenia revealed that visual, but not auditory dysfunction at age four was predictive of schizophrenia in adulthood (Schubert, Henriksson, & McNeil, 2005). Schizophrenia has been associated with impaired performance on a range of visual discrimination and recognition tasks, such as patterns, locations, movement trajectories and spatial frequency (O’Donnell et al., 1996), contrast sensitivity (Kéri, Antal, Szekeres, Benedek, & Janka, 2002; Slaghuis, 1998), and lower susceptibility to visual illusions compared to healthy controls (Dakin, Carlin, & Hemsley, 2005; King, Hodgekins, Chouinard, Chouinard, & Sperandio, 2017). Visual perceptual deficits were reported to be more predictive of conversion from a psychosis prodrome to schizophrenia than symptoms of thought and language problems and ideas of reference (Klosterkötter, Hellmich, Steinmeyer, & Schultze-Lutter, 2001), and are linked to functional outcome in schizophrenia (Green, Hellemann, Horan, Lee, & Wynn, 2012; Rassovsky, Horan, Lee, Sergi, & Green, 2011). Impaired visual processing has also been linked to transdiagnostic vulnerability in mental illness, often termed P or general psychopathology (Caspi et al., 2014). Recent studies have reported that individual differences in P are linked to aberrant structure (Romer et al., 2019) and functional connectivity of visual cortical areas of the brain (Elliott, Romer, Knodt, & Hariri, 2018).This underscores the clinical relevance of assessing early and low-level perceptual processes in severe mental illness.

Cognitive impairments are often reported among patients with schizophrenia (Kar & Jain, 2016), and are also found in bipolar disorder (Bourne et al., 2013; Simonsen et al., 2011). At the group level, patients with schizophrenia show reduced performance across several cognitive domains, including speed of processing, attention, vigilance, working memory, verbal learning and memory, visual learning and memory, reasoning and problem solving and verbal comprehension (Hartberg et al., 2011; Holmén et al., 2012; Nuechterlein et al., 2004; Vaskinn et al., 2011). Perceptual disruptions have been attributed to impaired executive functions such as flexible shifting of attention, inhibitory control and working memory (Jenkins, Bodapati, Sharma, & Rosen, 2018; Waters et al., 2012), and aberrant early sensory and perceptual processing may contribute to the clinical and cognitive symptoms in schizophrenia.

Here, using a computerized discrimination threshold estimation task along the basic visual sensory dimension of spatial frequency, we compared visual discrimination performance in patients with schizophrenia spectrum or bipolar disorders to healthy controls. In addition to testing for case-control differences, we investigated associations between discrimination threshold and positive and negative symptoms and IQ. Based on current theories and the reviewed literature we hypothesized that patients with severe mental disorders would exhibit higher discrimination thresholds, indicating worse performance, compared to healthy controls. Our final hypothesis is based on the notion of a hierarchical processing stream in which early sensory mechanisms represent precursors to more abstract and complex functions. We anticipated that higher visual discrimination threshold would be associated with more severe clinical symptoms as measured using the positive and negative syndrome scale (PANSS) and lower IQ.

## Materials and Methods

### Sample and exclusion criteria

A total of 165 healthy controls (HC) and 83 patients with severe mental disorders completed the task, including schizophrenia (n=28), schizophreniform (n=1), schizoaffective disorder (n=4), bipolar I disorder (n=25) and bipolar II disorder (n=30). Patients with schizophrenia, schizophreniform or schizoaffective disorders were combined into a schizophrenia spectrum disorders group (SZ, n=33), and patients with bipolar disorder I or bipolar disorder II were combined into a bipolar spectrum disorders group (BD, n=55). After exclusion based on task performance (see below and Supplemental Figure 1), 32 patients with SZ, 54 patients with BD and 148 HC were included in the analysis. Table 1 summarizes demographics and clinical variables. We assessed covariate balance by estimating standardized mean differences (Zhang, Kim, Lonjon, & Zhu, 2019) for SZ and BD versus HC, yielding the following results: SZ: IQ=-0.91, age= -0.71, sex=0.05; BD: IQ =-0.26, age=-0.45, sex=0.46. Distribution of age, IQ and education are shown in Supplemental Figure 2.

**Table 1.** Demographics and clinical characteristics including the subscales and total symptom score from PANSS for schizophrenia spectrum disorders (SZ), bipolar disorder (BD), and healthy controls, (HC). The Intelligence Quotient (IQ) was estimated from the Wechsler Abbreviated Scale of Intelligence (WASI).

|  | **HC** | **BD** | **SZ** |
| --- | --- | --- | --- |
| N | 148 | 54 | 32 |
| Age Mean (SD) years | 39.9 (9.6) | 33.8 (13.1) | 33.7 (8.54) |
| Sex (% female) | 44.5 | 64.8 | 50 |
| Education Mean (SD) years | 14.9 (2.1) | 14.5 (2.24) | 13.7 (2.71) |
| Discrimination threshold Mean (SD) | 0.11 (0.07) | 0.12 (0.07) | 0.14 (0.08) |
| PANSS Positive Mean (SD) | - | 9.04 (3.04) | 14.7 (4.89) |
| PANSS Negative Mean (SD) | - | 9.52 (2.93) | 14.0 (6.24) |
| PANSS General Mean (SD) | - | 25.7 (5.47) | 27.2 (6.42) |
| PANSS Total Mean (SD) | - | 44.3 (9.54) | 49.5 (13.7) |
| IQ Mean (SD) | 116 (9.06) | 113 (9.08) | 104 (12.6) |

Patients were recruited from psychiatric hospitals and outpatient clinics in the Oslo region, Norway, as part of the ongoing Thematically Organized Psychosis (TOP) study. Healthy controls from the same geographic area were identified by a stratified random draw from the national population registry and invited by letter. They were screened and excluded if they had experienced any psychiatric disorder, or their first-degree relatives had experienced a severe psychiatric disorder. Exclusion criteria included age => 18 years, IQ < 70, previous moderate or severe head injury or a neurologic illness. All participants gave written informed consent before participating. The study was approved by the Regional Committee of Medical Research Ethics and the Norwegian Data Inspectorate and was conducted in accordance with the Helsinki declaration.

### Clinical and cognitive assessment

Diagnoses were assessed by clinical psychologists or physicians using the Structured Clinical Interview for DSM-IV axis I disorders (SCID-I) modules A-E (First, Spitzer, Gibbon, & Williams, 1997). Diagnostic reliability has previously been found to be satisfactory (Ringen et al., 2008). Patients meeting criteria for DSM-IV axis diagnoses of schizophrenia spectrum disorders (schizophrenia, schizoaffective, schizophreniform) or bipolar disorder (Bipolar type I or II) were included, thus excluding participants diagnosed with non-specified mental disorders (including single-episode psychotic disorders, non-specified affective disorders).

Current psychotic symptoms were assessed with the Positive and Negative Syndrome Scale (PANSS; Kay, Fiszbein, & Opler, 1987), which is widely used in clinical and research settings and considered a reliable means of symptom assessment (Bell, Milstein, Beam-Goulet, Lysaker, & Cicchetti, 1992; Müller et al., 1998). Positive symptoms refer to an excess or distortion of normal functions (e.g. hallucinations and delusions), while negative symptoms represent a diminution or loss of normal functions. Estimated IQ scores were assessed using the Matrix Reasoning and the Vocabulary subtests from the Wechsler Abbreviated Scale of Intelligence, WASI (Wechsler, 2007).

Participants were excluded based of the following criteria: an estimated threshold >100% difference on any of the four rounds. Consequently, six participants were excluded (4 HC, 1 patient with SZ, 1 patient with BD). Supplemental Figure 1 shows a flow chart of the exclusion and inclusion of participants.

### Stimulus presentation

Figure 1 illustrates the experimental paradigm. The task was adapted from Sneve, Alnæs, Endestad, Greenlee, and Magnussen (2011), and consisted of sinusoidal grating patterns with varying spatial frequency. Each round of the task consisted of 32 trials, and each trial consisted of 0.5 second presentation of a sample grating, followed by a 1 second inter-stimulus interval (ISI), and then a 0.5 second presentation of the test grating, after which the participants indicated whether the test had a higher or lower spatial frequency compared to the sample by pressing the up or down-arrow on the keyboard (2-alternative forced choice). Each participant completed 4 rounds of the task. Participants were seated approximately 50-60 cm from the laptop screen (screen width 30.5 cm). The sinusoid had a maximum Michelson’s contrast of 0.6, tapered with a Gaussian kernel with a standard deviation of 2.5. Base cycles per visual degrees were set to vary between 2, 3 and 4 across trials, while orientations of the spatial frequency grids varied with 90 degrees across trials.

**Figure 1.**
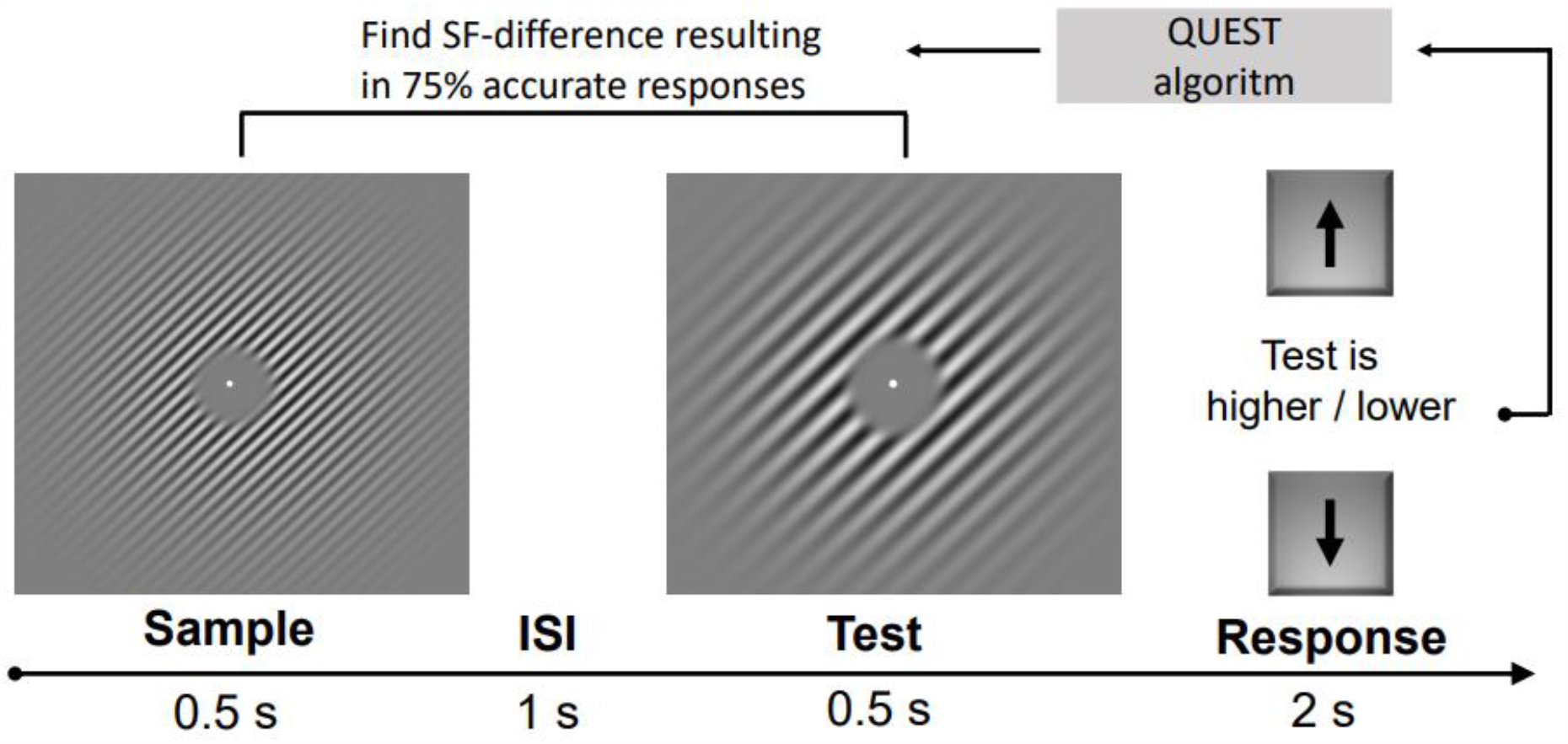
The visual discrimination task. Participants performed 4 rounds of a 2-alternative forced choice task where they indicated whether the test grating had a higher or lower spatial frequency compared to the sample stimulus, briefly interrupted by an inter-stimulus interval (ISI)

### Visual discrimination threshold estimation

The paradigm was coded and run using Psychtoolbox-3 (Kleiner, Brainard, & Pelli, 2007) in MATLAB (version 2015a; Mathworks, Natick, Massachusetts). Individual 75% discrimination thresholds were estimated using an adaptive maximum likelihood procedure (QUEST; Watson & Pelli, 1983). The QUEST algorithm varied the spatial frequency difference of the test stimulus compared to the sample stimulus, converging on the difference producing a discrimination accuracy of 75%. All participants completed 4 rounds of 32 trials. Each round took approximately 2 minutes to complete. The first estimation round always started using the same spatial frequency difference across participants, while subsequent runs used the previous estimated difference threshold as the updated initial value. Supplemental Figure 3 shows the bivariate correlation of estimated thresholds between the four rounds. Individual average threshold levels were then calculated as the average across the final three rounds.

### Statistical analyses

Statistical analyses were performed using R (version 3.5.1, https://www.r-project.org). Bivariate correlational analyses were performed to investigate whether threshold estimations converged across sessions. For group comparisons we employed robust linear regression (Threshold ∼ Diagnosis + Age + Sex, using “lmr” from the R package ‘MASS’ (Venables & Ripley, 2002), followed by a robust F-test (“f.robustftest” from the R package ‘sfsmisc’ (Maechler et al., 2021) and post-hoc comparisons of group differences corrected using Tukey (using “glht” from the R-package ‘multcomp’(Holthorn, Bretz, & Westfall, 2008). Robust regression weights data points to reduce the impact of outliers and noisy observations, while still including all the data in the analysis. For transparency, we also report the results from a standard linear regression and the accompanying post-hoc tests

Finally, we tested for associations between visual discrimination threshold and PANSS domain scores and IQ using separate multiple linear regression analyses within groups (PANSS only available from patients), with age and sex as covariates (*Threshold ∼ Age + Sex + Cognition/Clinical*). Clinical symptom scores were calculated using PANSS subscales, separating positive, negative and generalized symptoms (Kay et al., 1987; Van den Oord et al., 2006). Compared to healthy controls, IQ was significantly lower among patients with schizophrenia spectrum disorder (t=-5.36, p=.5.3e-08), but not among patients with bipolar disorder (t=-1.11, p=.27). Since our design does not allow us to disentangle the effects of IQ from diagnosis, we investigated associations between visual discrimination performance and IQ within groups.

## Results

### Group differences in visual discrimination threshold

Figure 2 shows the distribution of estimated threshold by group. Robust regression revealed a significant effect of diagnosis on discrimination threshold (robust F=6.76, p=.001). Post-hoc comparisons corrected for multiple comparisons using Tukey revealed that patients with SZ (mean = 14%, SD = 0.08) had significantly higher estimated discrimination thresholds than healthy controls (mean=10.8%, SD = 0.07, β = 0.35, t=3.4, p=0.002), as did patients with BD (mean = 12.3%, SD = 0.07, β= 0.21, t=2.42, p=0.04). There was no significant difference between BD and SZ (β = -0.14, t=-1.2, p=0.45).

**Figure 2.**
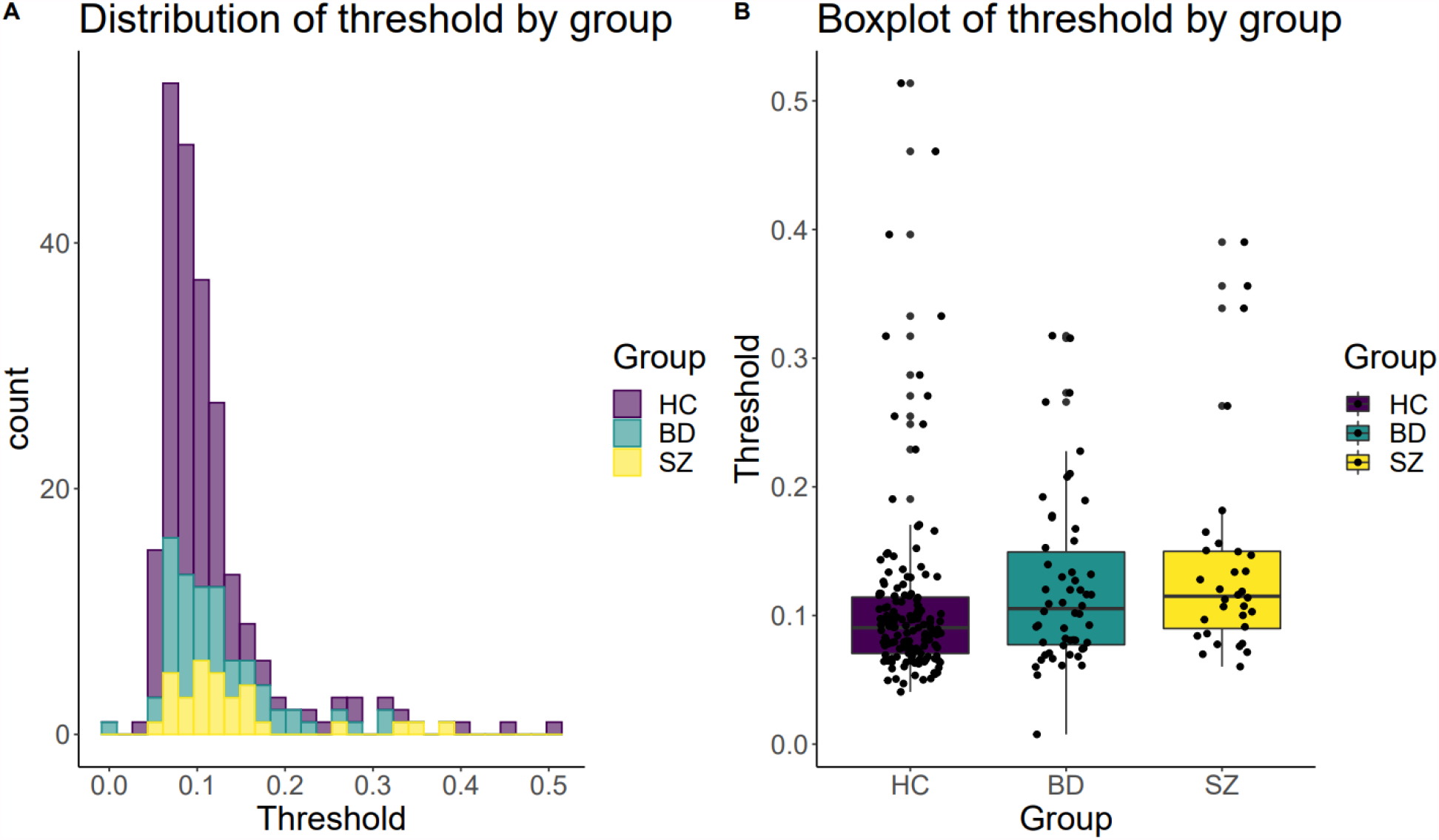
A) The distribution of visual discrimination thresholds by diagnostic group. B) Boxplot of visual discrimination threshold by diagnostic group. Robust regression revealed significant differences in threshold for both schizophrenia (SZ) and bipolar (BD) patients vs healthy controls (HC), while no difference was found for SZ vs BD.

The standard linear regression (Threshold ∼ Diagnosis + Age + Sex) showed a significant main effect of diagnosis (F=3.6, p=.03). Post-hoc pairwise group comparisons corrected for multiple comparisons using Tukey revealed significantly higher threshold among patients with SZ compared to HC (β=0.49, t=2.5, p=.03), and no significant differences between patients with BD HC (β=0.25 t=1.56, p=.26), nor between the two patient groups (β=-0.24, t=-1.09, p=.52).

### Associations between visual discrimination performance and IQ

Figure 3A shows the relationship between discrimination threshold and IQ. Linear models across all participants revealed a significant negative association between visual discrimination threshold and IQ when controlling for diagnostic group (β = -0.3, t=-3.43, p=0.0007). Within groups, a significant negative association was found among healthy controls (β=-0.29, t=-3.72, p=.0003) and patients with bipolar disorder (β=-0.34, t=-2.53, p=.015), but not among patients with schizophrenia spectrum disorder (β=-0.22, t=-1.34, p=.19). Follow-up analysis revealed no significant interaction effect for diagnostic group on IQ (F = 0.044, p=.97).

**Figure 3.**
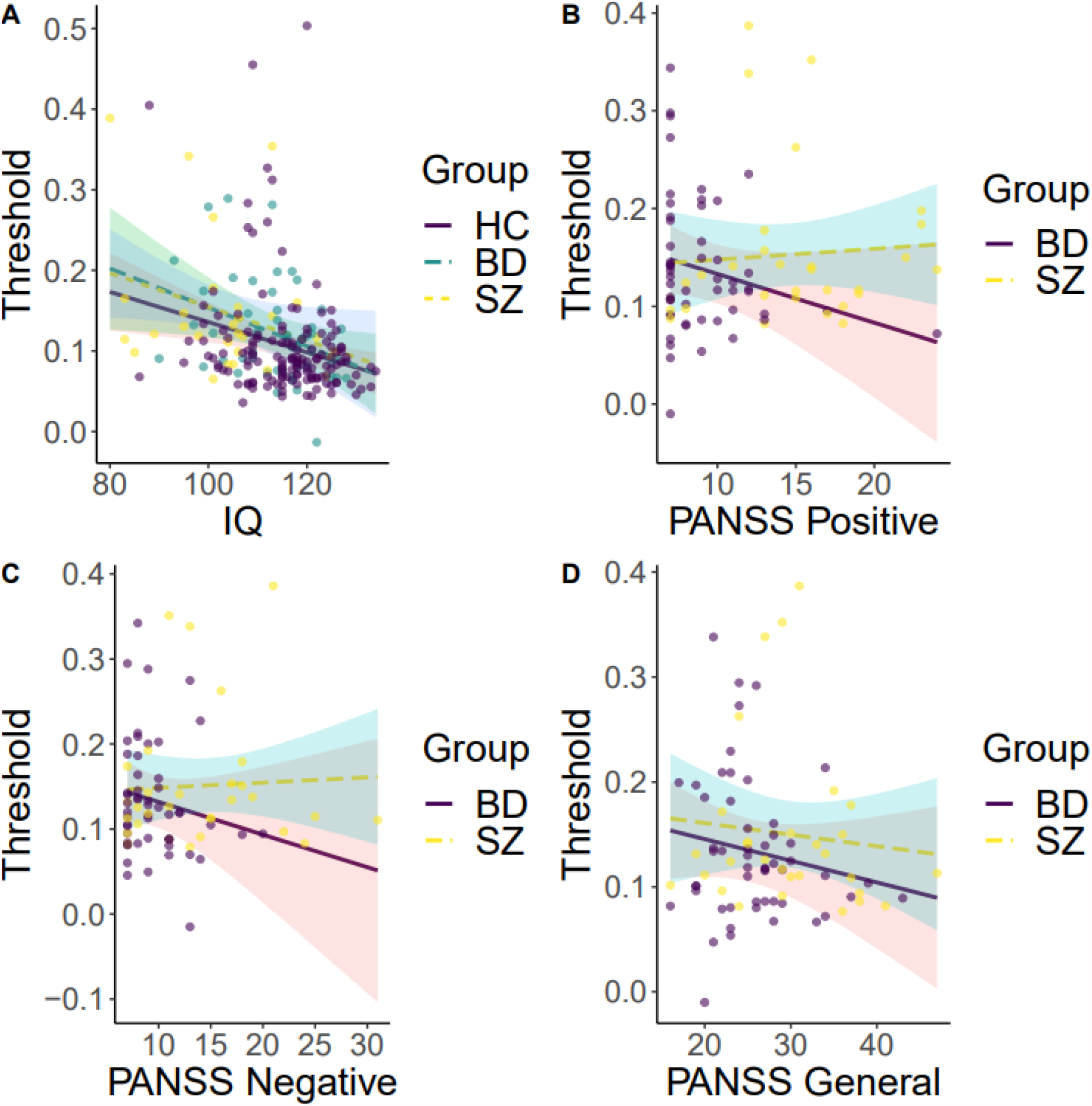
There was no significant IQ by group interaction. For within-group analysis, IQ showed a significant association with discrimination threshold. There were no significant associations of any of the PANSS scores with discrimination threshold. **A)** Discrimination thresholds plotted against IQ, by group. **B**) Discrimination thresholds plotted against positive PANSS scores, by diagnosis **C**) Discrimination thresholds plotted against negative PANSS scores, by diagnosis. **D**) Discrimination thresholds plotted against general PANSS scores, by diagnosis.

### Associations between visual discrimination performance and PANSS symptom domains

Figures 3B-D depict the associations between PANSS domain scores and visual discrimination threshold. Linear models within patients with SZ revealed no significant association between discrimination threshold and the PANSS positive (β=0.104, t=0.58, p=.6), negative (β=-0.01, t=0.046, p=.96) and general (β=-0.08, t=-0.5, p=.64) subscales. For patients with BD, no significant associations were found for the PANSS positive (β=-0.24, t=-1.222, p=.227), negative (β=-0.23, t=-1.09, p=.281), nor the general (β=-0.21, t=-1.37, p=.178) subscales.

## Discussion

Patients with severe mental illnesses including schizophrenia spectrum and bipolar disorders often experience neurocognitive deficits across several domains. Here, we show that patients with schizophrenia spectrum or bipolar disorders have higher visual discrimination thresholds for the basic visual feature of spatial frequencies compared to healthy controls. This is consistent with previous reports of basic visual processing deficits in patients with severe mental disorders (Brenner, Lysaker, Wilt, & O’Donnell, 2002; Silverstein & Rosen, 2015). Overall, the results support the notion of disrupted basic visual processing in patients with schizophrenia (Butler et al., 2005; Li, 2002; Slaghuis, 1998), and are in line with previous reports of visual discrimination deficits in patients with bipolar disorder (O’Bryan et al., 2014).

Patients with schizophrenia and bipolar disorders are heterogeneous both at the clinical, cognitive and brain biological level (Alnæs et al., 2019; Joyce & Roiser, 2007; Wolfers et al., 2018; Wolfers et al., 2021). Some of this clinical heterogeneity may be resolved by combining clinical profiling with detailed neurocognitive assessment across the hierarchical processing streams from early sensory and perceptual mechanisms to later and more abstract processes including decision making and reasoning. In the case of the current study, we found no association with the level of psychotic symptoms.

Disrupted early visual processing in patients with severe mental illness has also been linked to dysfunction in neurotransmission systems (Javitt, 2009b), with reduced visual cortical GABA concentration associated with aberrant surround suppression in patients with schizophrenia (Yoon et al., 2010; Yoon et al., 2009). Taken together, these studies suggest that early visual processing impairments could be linked to the underlying vulnerability for mental illness, and possibly serve as a proxy of impaired basic brain functions, rather than simply resulting from higher-order deficits of attention and executive function (Sponheim, Sass, Noukki, & Hegeman, 2013). Studies investigating the role of visual perceptual processing and associations with general risk factors for mental illness in childhood and adolescence are therefore warranted.

Our analysis revealed negative associations between IQ and visual discrimination thresholds across all participants, and specifically within HC and BD patients. Power-limitations may explain why we couldn’t establish this relationship in our SZ sample, as early visual processing as precursors to higher-order cognitive functioning are frequently reported in the literature (Acton & Schroeder, 2001; Arranz-Paraíso & Serrano-Pedraza, 2018; Cook, Hammett, & Larsson, 2016; Melnick, Harrison, Park, Bennetto, & Tadin, 2013), and the notion of early visual processing as precursors of higher-order cognitive functioning. Our results did not show any significant associations between discrimination thresholds and symptom severity. There is always the possibility that we were not able to detect a true relationship due to the relatively small variation in PANSS scores in our sample, resulting in a type II error. Previous studies have reported associations between impaired sensory discrimination and poor cognitive functioning in people with psychosis that are not solely explained by their psychiatric symptoms (Ramsay et al., 2020). It has been suggested that sensory disruptions, for example in motion processing, are due to higher-order memory processing deficits, caused or influenced by existing disruptions in top-down processes (Brenner, Wilt, Lysaker, Koyfman, & O’Donnell, 2003). Future studies may be able to investigate the integration between such top-down and bottom-up processes and its relation to the observed cognitive deficits in patients with severe mental disorders.

Some studies have reported associations between symptom severity measured with the PANSS and performance on an auditory tone matching task, where the mismatch-negativity (MMN) as measured using EEG (Javitt, Shelley, & Ritter, 2000) was inversely associated with right-ear performance on a dichotic listening task (Hugdahl et al., 2012) and performance on a face recognition task (Chen, Norton, McBain, Ongur, & Heckers, 2009). In contrast, our analysis revealed no significant associations between PANSS domain scores and visual discrimination thresholds, and our findings are thus in line with studies reporting no robust associations between visual performance and symptom load (Aleman, Böcker, Hijman, de Haan, & Kahn, 2003; Brenner et al., 2002).

Of interest to future studies and potential for clinical implementation, performance on sensory discrimination is prone to training effects (Adini, Wilkonsky, Haspel, Tsodyks, & Sagi, 2004), termed perceptual plasticity, and robust plasticity has been reported for the visual system in patients with schizophrenia (Norton, McBain, Öngür, & Chen, 2011). This could in principle be used to develop bottom-up behavioral strategies as a part of an intervention or treatment protocol. Future investigations may also assess the relationships between sensory discrimination deficits across auditory and visual modalities, as auditory perceptual disruptions are reported as more frequent. Previous studies have suggested a link between the two (Ramsay et al., 2020).

The current findings should be interpreted in light of some limitations. Effects of medication were not explicitly tested for in this study. Previous studies have suggested that medication may partly explain mixed results on behavioral, perceptual and cognitive deficits among patients with severe mental illness. Our design did not allow us to rule out medication effects on visual discrimination ability (Fernandes et al., 2019). Investigating effects of medication requires experimental control of medication type and doses, and was therefore not feasible in the current study. The current experimental task was employed as part of a brief assessment prior to brain MRI assessment. While demonstrating feasibility in a clinical setting, stricter experimental control of variables such as ambient light, visual acuity and distance to screen as afforded by a psychophysics lab environment would also be beneficial for future studies employing similar psychophysical methods. The interval of 1 second between sample and test stimuli in the paradigm makes it difficult to disentangling differential disruptions in encoding and retention of stimuli, reflecting separate but interacting processes in visual working memory, involving overlapping neural systems (Pasternak & Greenlee, 2005).

Also, patient sample sizes and thereby statistical power could have affected our ability to probe the relationship between visual perception and clinical symptoms. Cognitive deficits in bipolar disorders are frequently reported, but typically less severe than those observed for schizophrenia (Daban et al., 2006). All our groups had mean IQ levels which are nominally larger than the population average, indicating that our participants represent a relatively cognitive high-functioning part of the population, which is of relevance for the generalizability of our results to the clinical and healthy population at large. Future studies may investigate a broader range of the clinical severity spectrum. Further, the difference between patients with bipolar disorder and healthy controls was significant using robust linear regression, but not when using a standard linear model, suggesting outliers may mask group differences.

In conclusion, our findings of early visual processing deficits in patients diagnosed with schizophrenia spectrum or bipolar disorders, suggest an association between early visual processing and general intellectual abilities in patients with bipolar disorder and healthy controls. Taken together, these findings elucidate underlying disease mechanisms in severe mental illness. Future studies investigating brain mechanisms associated with such basic sensory deficits, including functional brain imaging and mapping of neurotransmitter function in individual patients have the potential to yield important insights into the neurocognitive mechanisms of psychotic disorders.

## Data Availability

Open science and data availability statement:
Relevant scripts and scrambled and anonymized data needed to reconstruct the reported analyses and results is available on OSF (link below). While the sensitive nature of the data and our current approvals do not allow for public sharing of real data, anonymized data may be available upon request to the corresponding author following appropriate data transfer agreements.
OSF-repository: https://osf.io/r64td/?view_only=8bee034ade5a46dc91b0e95654c2e780.

https://osf.io/r64td/?view_only=8bee034ade5a46dc91b0e95654c2e780

## Acknowledgements

A. R. L is funded by a student grant awarded from the Research Council of Norway (2018/14182). D.A. is funded by the South-Eastern Norway Regional Health Authority (2019107, 2020086) and Bjørknes College. K. K. K is funded by the South-East Norway Regional Health Authority (2015044). AM.G.dL. is funded by the Swiss National Science Foundation (PZ00P3_193658). M.H.S. is funded by Department of Psychology, University of Oslo. T.V.L. is funded by Oslo University Hospital. I.M is funded by NORMENT, University of Oslo. O.A.A. is funded by the Research Council of Norway (223273, 283799, 2837989), the South-East Norway Regional Health Authority (2019-108) and KG Jebsen Stiftelsen. L.T.W. is funded by the European Research Council under the European Union’s Horizon 2020 research and innovation program (ERC Starting Grant 802998), the Research Council of Norway (249795, 300768), the South-East Norway Regional Health Authority (2019101), the Research Council of Norway (298646, 300767), Division of Mental Health and Addiction, Oslo University Hospital and Department of Psychology, University of Oslo.

## Open science and data availability statement

Relevant scripts and scrambled and anonymized data needed to reconstruct the reported analyses and results is available on OSF (link below). While the sensitive nature of the data and our current approvals do not allow for public sharing of real data, anonymized data may be available upon request to the corresponding author following appropriate data transfer agreements. OSF-repository: https://osf.io/r64td/?view_only=8bee034ade5a46dc91b0e95654c2e780.

