## Supplemental figures for "Spatial frequency discrimination in patients with schizophrenia spectrum and bipolar disorders: Evidence of early visual processing deficits and associations with intellectual abilities"

### Supplementary figures

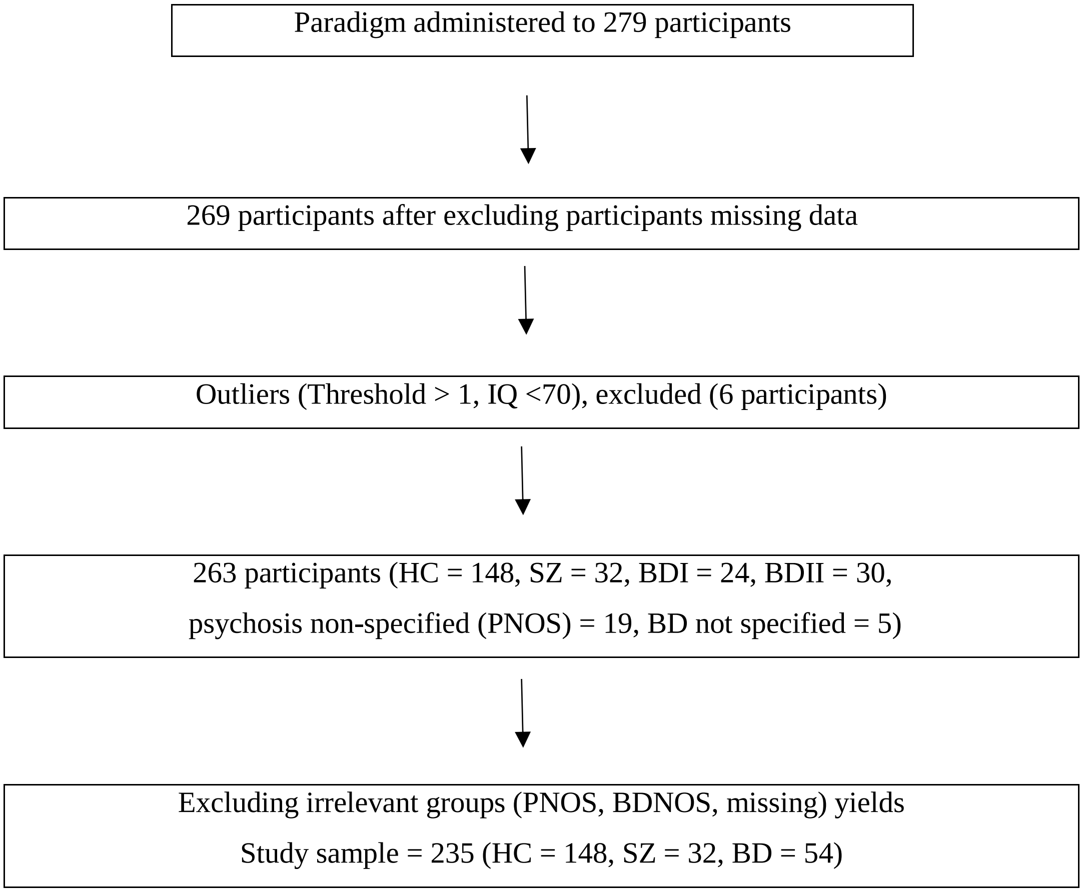


**Supplemental Figure 1.** Flow chart showing inclusion and exclusion of study sample.

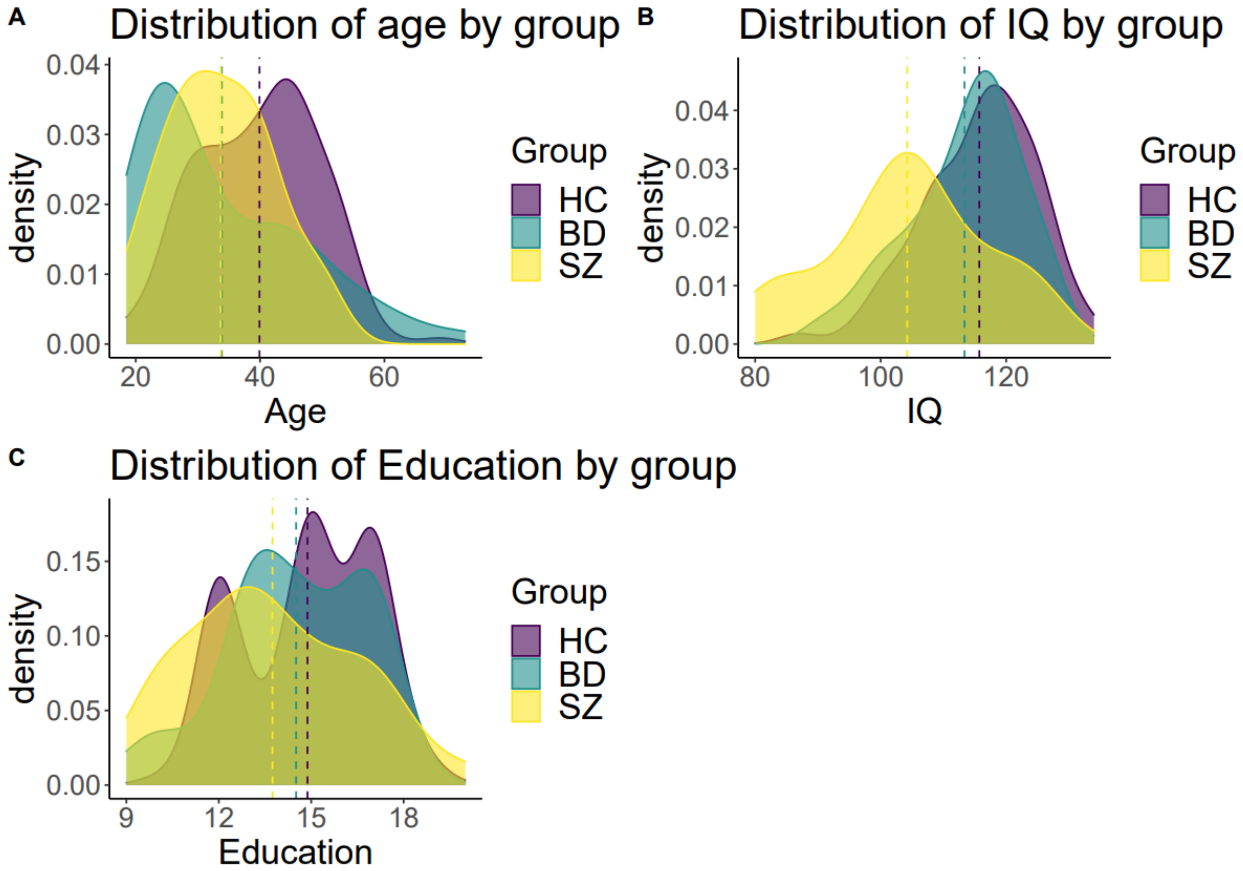


**Supplemental Figure 2.** A: Distribution of age by group, B: Distribution of IQ by group, C: Distribution of Education by group

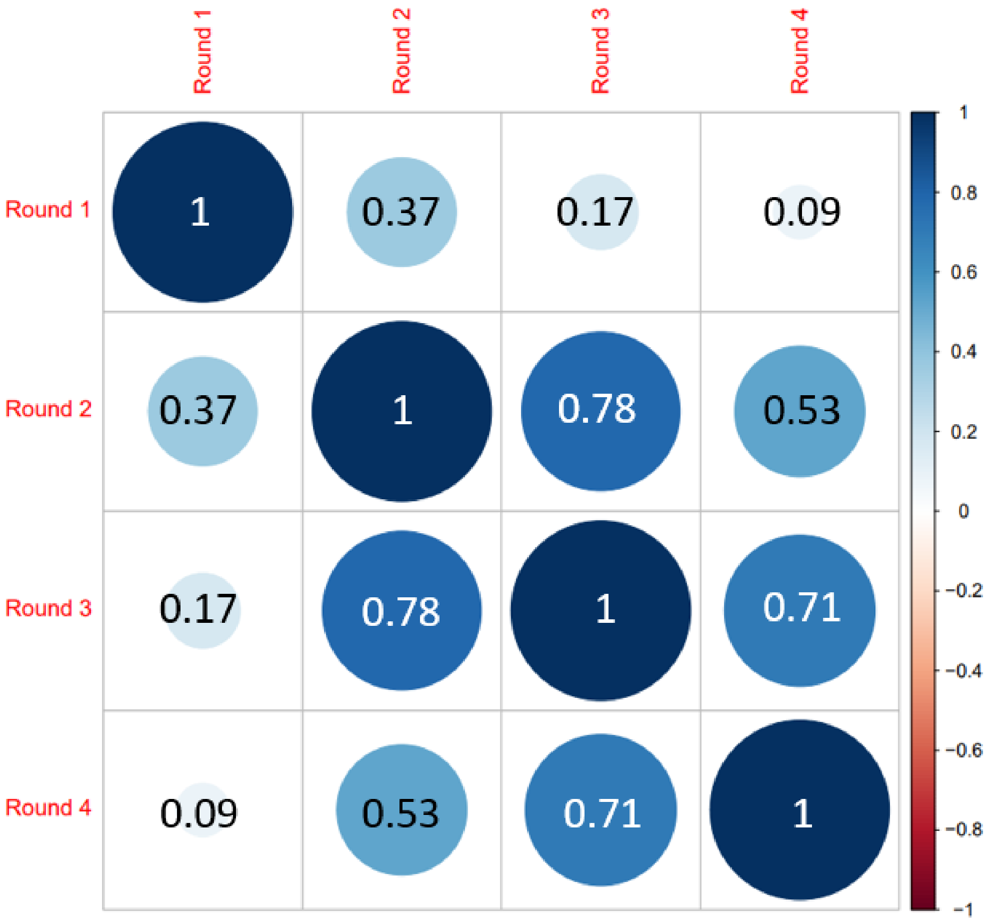


**Supplemental Figure 3.** Bivariate correlational analyses of estimated thresholds across the four rounds. As expected, correlations are higher for the latest compared to the first rounds, since participants started at the same initial threshold, before the algorithm converged on each individual's threshold toward the later rounds.
